# Non-invasive Temporal Interference Stimulation of the Hippocampus Suppresses Epileptic Biomarkers in Patients with Epilepsy: Biophysical Differences between Kilohertz and Amplitude Modulated Stimulation

**DOI:** 10.1101/2024.12.05.24303799

**Authors:** Florian Missey, Emma Acerbo, Adam S. Dickey, Jan Trajlinek, Ondřej Studnička, Claudia Lubrano, Mariane de Araújo e Silva, Evan Brady, Vit Všianský, Johanna Szabo, Irena Dolezalova, Daniel Fabo, Martin Pail, Claire-Anne Gutekunst, Rosanna Migliore, Michele Migliore, Stanislas Lagarde, Romain Carron, Fariba Karimi, Raul Castillo Astorga, Antonino M. Cassara, Niels Kuster, Esra Neufeld, Fabrice Bartolomei, Nigel P. Pedersen, Robert E. Gross, Viktor Jirsa, Daniel L. Drane, Milan Brázdil, Adam Williamson

**Affiliations:** International Clinical Research Center, St. Anne’s University Hospital Brno, 60200 Brno, Czech Republic; Aix-Marseille Université́, Inserm, Institut de Neurosciences des Systèmes (INS) UMR_S 1106, Marseille, France; Department of Neurosurgery, Emory University School of Medicine, Atlanta, Georgia, USA; Department of Neurology, Emory University School of Medicine, Atlanta, Georgia, USA; Brno Epilepsy Center, 1st Department of Neurology, St. Anne’s Univ. Hospital and Faculty of Medicine, Masaryk University, member of the ERN EpiCARE. 60200 Brno, Czech Republic; Institute of Neurosurgery and Neurointervention, Semmelweis University, Budapest, Hungary; Institute of Biophysics, National Research Council, Palermo, Italy; SUNY Dept. of Neurology, Downstate Health Science University, Brooklyn, NY, USA; APHM, Timone Hospital, Epileptology Department, Marseille, France; Department of Functional and Stereotactic Neurosurgery, Timone University Hospital, Marseille, France; Foundation for Research on Information Technologies in Society (IT’IS), Zurich, Switzerland; Department of Information Technology and Electrical Engineering, Swiss Federal Institute of Technology (ETH), Zurich, Switzerland; Department of Neurology, School of Medicine and Center for Neuroscience, University of California, Davis, CA USA; Department of Neurosurgery, New Jersey Medical School and Robert Wood Johnson Medical School, Rutgers University, New Jersey, USA; Departments of Pediatrics, Emory University School of Medicine, Atlanta, GA USA; Department of Neurology, University of Washington School of Medicine, Seattle, WA USA; Center for Social and Affective Neuroscience, Department of Biomedical and Clinical Sciences, Linköping University, Sweden

**Keywords:** Temporally Interfering Electric Fields, Non-Invasive Brain Stimulation, Neuromodulation, Epileptic Biomarkers, sEEG, Amplitude Modulation, Conduction Block

## Abstract

Medication-refractory focal epilepsy creates a significant challenge, with approximately 30% of patients ineligible for surgery due to the involvement of eloquent cortex in the epileptogenic network. For such patients with limited surgical options, electrical neuromodulation represents a promising alternative therapy. In this study, we investigate the potential of non-invasive temporal interference (TI) electrical stimulation to reduce epileptic biomarkers in patients with epilepsy by comparing intracerebral recordings obtained before, during, and after TI stimulation, to recordings during low and high kHz frequency (HF) sham stimulation. Thirteen patients with symptoms of mesiotemporal epilepsy (MTLE) and implanted with stereoelectroencephalography (sEEG) depth electrodes received TI stimulation with an amplitude modulation (AM) frequency of 130Hz (Δf), where the AM was delivered with lower frequency kHz carriers (1kHz + 1.13kHz), or higher frequency carriers (9kHz + 9.13kHz), targeting the hippocampus – a common epileptic focus and consequently stimulation target in MTLE. Our results show that TI stimulation yields a statistically significant decrease in interictal epileptiform discharges (IEDs) and pathological high-frequency oscillations (HFOs) – specifically fast-ripples (FR) –, where the suppression is apparent in the hippocampal focus and propagation from the focus is reduced brain-wide. HF sham stimulation at 1kHz frequency also impacted the IED rate in the cortex, but without reaching the hippocampal focus. The HF sham effect diminished with increasing frequencies (2, 5, and 9kHz, respectively), specifically as a function of depth into the cortex. This depth dependence was not observed with the TI, independent of the employed carrier frequency (low or high kHz). Furthermore, a strong carry-over effect, i.e., suppression of epileptic biomarkers for a period of time after the end of stimulation, was observed for TI but not for kHz. Our findings underscore the possible application of TI in epilepsy, as an additional non-invasive brain stimulation tool, potentially offering opportunities to assess brain region response to electrical neuromodulation before committing to a deep brain stimulation (DBS) or responsive neurostimulation (RNS) implants. Our results further demonstrate distinct biophysical differences between kHz and focal AM stimulation.

**Graphical Abstract:** 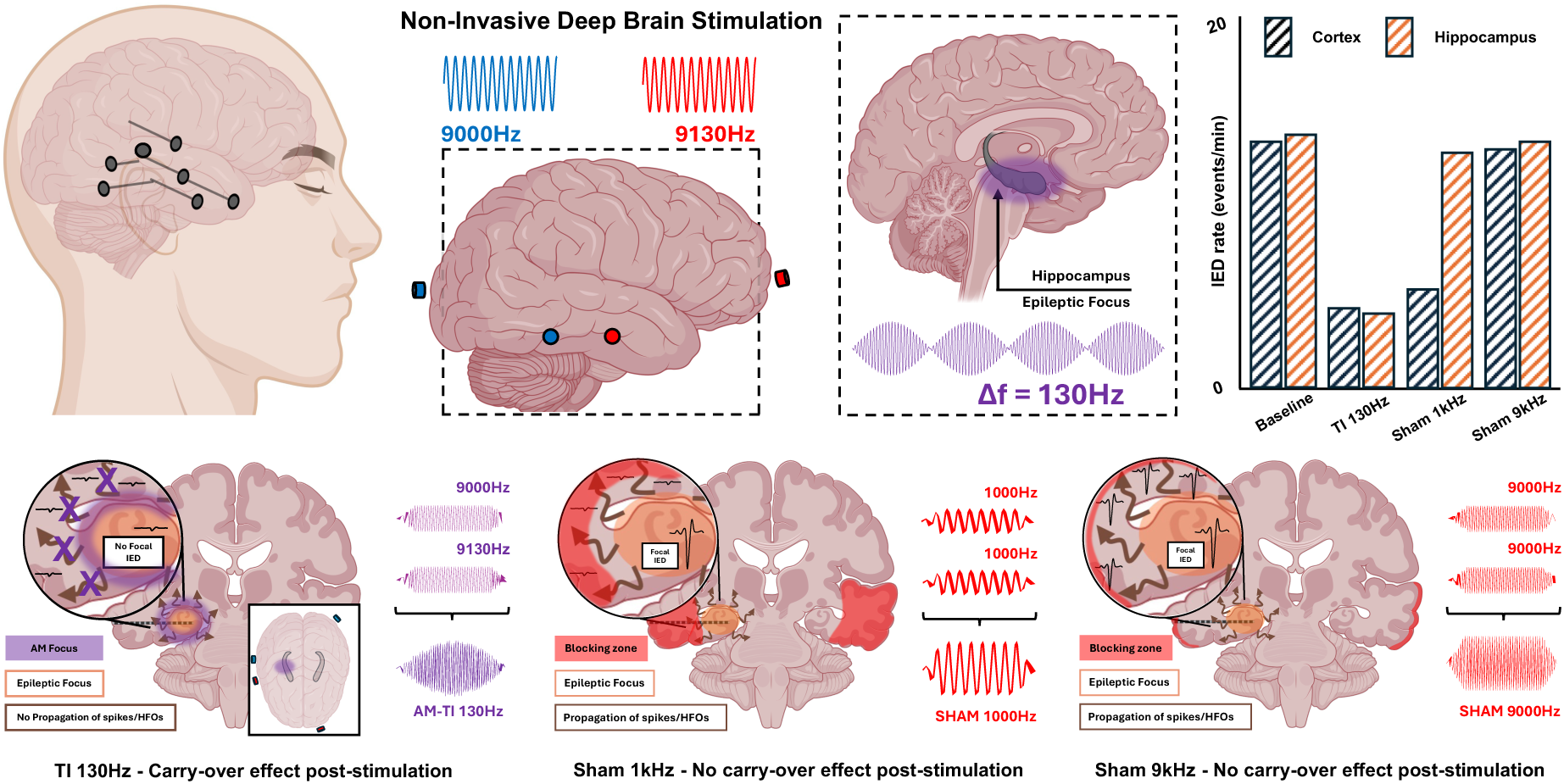

## Introduction

Epilepsy presents a significant neurological challenge, as the origins of seizure generation in the brain are highly patient-specific, limiting initial treatment options to generalized medications which lack targeted precision^1^. Additionally, one-third of patients with seizures are drug-resistant, leaving resective surgery as the primary treatment option^2^. However, approximately 30% of drug-resistant patients are not suitable candidates for resective surgery due to the high functional importance of areas necessitating resection^2^. In such cases, invasive brain stimulation – specifically deep brain stimulation (DBS) or responsive neurostimulation (RNS) – is typically the remaining therapeutic option^3^. Alternative neuromodulation treatments, such as Vagus Nerve Stimulation (VNS), are available for drug-resistant epilepsies but generally do not achieve complete seizure freedom^4–7^.

Both DBS and RNS are challenging, as there are numerous potential targets (e.g., anterior nucleus of the thalamus - ANT, centromedian nucleus of the thalamus - CMT, pulvinar, hippocampus, and neocortex)^8–12^, and only a small number of targets (notably the ANT, CMT and hippocampus) have been thoroughly evaluated in double-blinded studies. DBS stimulation at 130-145Hz of either the hippocampus or ANT resulted in a reduction of seizure frequency^10–14^, along with a decrease in interictal epileptiform discharges (IEDs) in temporal lobe epilepsy patients^15,16,17,18^. A motivation for our study is that a subset of patients do not respond favorably to DBS or RNS and can suffer cognitive side effects, which are difficult to predict ahead of implantation^19–21^.

Non-invasive brain stimulation techniques targeting these regions identified as suitable DBS or RNS locations, could support the prediction of post-implant side effects prior to invasive implantation. The most common non-invasive techniques include transcranial alternating current stimulation (tACS), transcranial direct current stimulation (tDCS), and transcranial magnetic stimulation (TMS) – techniques with applications in both research and clinical practice^22^. The methods modulate brain activity via electric currents delivered through the scalp and skull, or induced magnetically, and influence neuronal excitability, connectivity, and plasticity, ultimately leading to changes in brain function^23^. However, efficacy of traditional non-invasive methods in the treatment of epilepsy is limited, and the methods are typically considered applicable only to shallower cortical targets and not to the deep structures associated with therapeutic invasive DBS^24^.

Temporal Interference (TI) stimulation is an emerging non-invasive electrical stimulation technique which allows electrical modulation of deep brain structures. Unlike traditional methods, TI applies high frequency currents (>1kHz) using a minimum of two independent pairs of transcutaneous stimulation electrodes. The employed frequencies differ slightly, resulting in an amplitude-modulated field because of alternating phases of constructive and destructive interference. The kHz current pathways are optimized to maximally and selectively amplitude-modulate the field at a specific deep brain target where the fields overlap^25^. The amplitude modulation (AM) frequency is equal to the frequency difference (Δf = |f1 – f2|). When Δf is in the physiological range, there is evidence that neural activity is modulated. Notably, the frequency of the AM in previous experiments has been selected to match conventionally applied DBS frequencies to produce similar effects^26^. TI has been tested in rodent^27–32^ and non-human primate^33^ models, and more recently, in healthy human subjects^34^.

We have previously employed TI stimulation using a 130 Hz envelope frequency, a frequency often used for invasive DBS in epilepsy patients and in epileptic animal models, and known to suppress epileptic biomarkers^27^.

In this work, we analyzed the impact of TI with a 130Hz AM signal in patients with epilepsy. Patients implanted with stereoelectroencephalography (sEEG) depth electrodes were hospitalized for 2 to 3 weeks to assess potential resective surgery targets. sEEG electrodes are implanted to record intracranial electrophysiological signals and stimulate precise deep brain areas in order to assist in the delineation of the epileptogenic zone (EZ) and its relation with eloquent cortices. Utilizing recordings from the sEEG electrodes during TI, we were able to investigate alterations in epileptic biomarkers as a function of TI stimulation (Δf=130Hz frequency modulation) and to map the applied AM signal in order to ascertain its hippocampal focality. The study has taken place at three research centers, Emory University (USA), St. Anne’s University Hospital (Czech Republic), and Semmelweis University (Hungary).

Our results demonstrate that TI stimulation significantly decreases interictal epileptiform discharges (IEDs) and pathological high-frequency oscillations (HFOs) – specifically fast-ripples (FR) – within the hippocampal focus and reduces propagation across the brain. In contrast, sham stimulation at lower kilohertz (kHz) frequencies impacted cortical but not hippocampal IEDs, with diminishing effectiveness at increasing kHz frequency. Furthermore, a therapeutic carrier-over effect – the suppression of epileptic biomarkers for a period of time after the end of stimulation – was only observed for AM and not for unmodulated kHz. The results suggest distinct differences in biophysical mechanisms and associated response characteristics from kHz compared to focal AM.

## Methods

*The study is registered as a clinical trial with clinicaltrials.gov (NCT06716866)*.

### Patients

13 patients with drug-resistant focal epilepsy and a clinical diagnosis of medial temporal epilepsy participated in the study after providing informed consent. All procedures involving human participants were conducted following the ethical standards of the institutional and/or national research committee (IRB00099109 Emory University, IIT/2023/25 Saint-Anne University Hospital - SAUH, OGYÉI/56526-2/2023 Institute of Neurosurgery and Neurointervention, Semmelweis University - INN-SU) and in accordance with the 1964 Helsinki Declaration and its later amendments or comparable ethical standards. All participants underwent video-EEG characterization of seizures and either 1.5 or 3 Tesla MRI, and PET scans in some patients. Presurgical non-invasive examinations, including high-resolution MRI scans, were performed to assess patient eligibility. Intracerebral multi-contact electrodes (Alcis® - INN-SU, Hungarian center; Dixi® - Emory University, USA center and SAUH, Czech center; 10-18 contacts) were surgically implanted for sEEG exploration. Postoperative computed tomography (CT) scans and/or MRI scans were conducted to verify the absence of complications and ensure accurate electrode placement using the GUI-based open-source application GARDEL^35^. Across all centers, patients underwent the stimulation protocols 6 to 10 days post-implantation.

### Modeling and simulations

Finite element simulations were executed utilizing both Sim4Life and the Sim4Life TI Planning Tool software developed by Zurich MedTech AG to estimate temporal interference stimulation. The simulations solved the ohmic-current-dominated electro-quasistatic equation ∇(σ∇ϕ) = 0. Here, σ denotes the local electrical conductivity, ϕ the electric potential, and the E-field is obtained as E=-∇ϕ. The ohmic-current-dominated electro-quasistatic approximation of Maxwell’s equations is suitable because σ≫ωɛ (ω: angular frequency, ɛ: permittivity, i.e., ohmic currents dominate over displacement currents) and the wavelength is much larger than the domain size.

The human model utilized in the Sim4Life simulations was derived from patient-specific MRI scans (co-registered T1 and CT). The head model for each patient included the associated implanted sEEG electrodes. All patients had the sEEG locations included in the model for postprocessing. However, only the patients from the FNUSA center had the sEEG electrodes properly modeled with recording contacts as Perfect Electrical Conductors (PEC) and inter-contacts as insulators. Tissue and electrode conductivities were automatically allocated to the model based on the ITI’S Foundation tissue properties database^36^ (low-frequency conductivities section). Stimulation electrodes mirrored the shape of the gel-based electrodes used in human experiments. Simulation results were normalized to total current, after applying Dirichlet boundary conditions at active electrodes. Equation 1 from Grossman et al., 2017^25^, was used to calculate the maximum modulation amplitude:

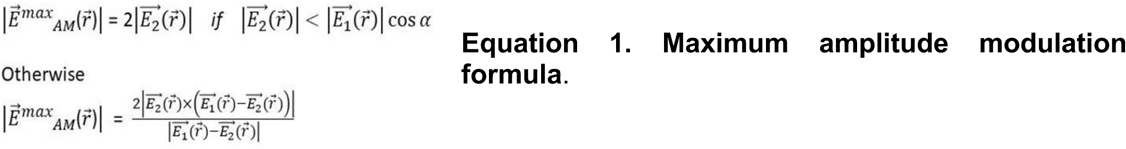

### Recordings

All studies were conducted with participants in the waking state. sEEG signals were recorded digitally (1024 or 2048 Hz, Natus Medical Incorporated®, Emory), or with a BioSDA09 (25 kHz, M&I, spol. s r.o.®, INN-SU and SAUH). The latter has an input signal voltage of +/− 25 mV, with an optional hardware filter of 0.01 Hz - 10 kHz), which allows to monitor stimulation voltages (sEEG artifact) during TI or sham stimulation.

### Stimulation (TI/sham)

Stimulation was applied to target the mesial temporal lobe with the target centered on the head of the hippocampus given that most IEDs originated from the hippocampus and the hippocampal formation was part of the epileptogenic network from which seizures originated.

TI stimulation was performed using two DS5 devices (Digitimer®, UK) driven by a function generator (Keysight®). Scalp electrodes (circular-shaped gel-assisted ECG electrodes, Ag/AgCl, 0.8 cm diameter, Ambu® or FIAB®) were used for TI stimulation. They were placed according to *Violante et al.*^34^ to target the hippocampus and in accord with Sim4Life modeling. Frequencies of TI stimulation varied among centers: Emory used 1300 and 1430 Hz, and the Czech and Hungarian centers 9000 and 9130 Hz, applying ±2 mA per pair (4 mA peak-to-peak). There were two protocols: In the first (SAUH and FNUSA), 20 minutes of baseline recording, followed by either sham stimulation (day one at SAUH and FNUSA), or active TI (day two), followed by 20 minutes of recording after the cessation of TI (see Figure 2A). At Emory University, all participants underwent 20 minutes of baseline, 20 minutes of sham/active TI and 20 minutes of post-TI recording (day one and two), with an additional recording of 20 minutes on day three. In the sham condition, the same carrier frequency was applied to both channels (Δf=0Hz, same current magnitudes as applied in the TI session), such that no amplitude modulation resulted. In the active TI condition, there was an offset frequency of 130 Hz - a typical frequency used for neuromodulation in epilepsy.

### AM Analysis

Gardel was used to accurately localize sEEG electrode contacts. Briefly, the T1 MRI was co-registered with the post-implantation CT and electrodes were labeled. For the analysis of TI waveforms, the recorded signal underwent bandpass filtering (1 kHz - 10 kHz). The AM magnitude was computed using the Hilbert transformation of the filtered signal. A sliding window of 230ms was employed to determine the peak-to-peak amplitude of the AM in mV by subtracting the minimum from the maximum AM values. The median value across all windows was utilized as the amplitude value for each contact. This process was conducted for each contact, providing amplitude values per contact (the reference was set as the averaged signal from all the electrodes in the brain). Finally, the amplitude values were projected onto the electrodes within the anatomical mesh of the patient to visually assess which brain regions received stimulation (Figure 1B).

**Figure 1.**
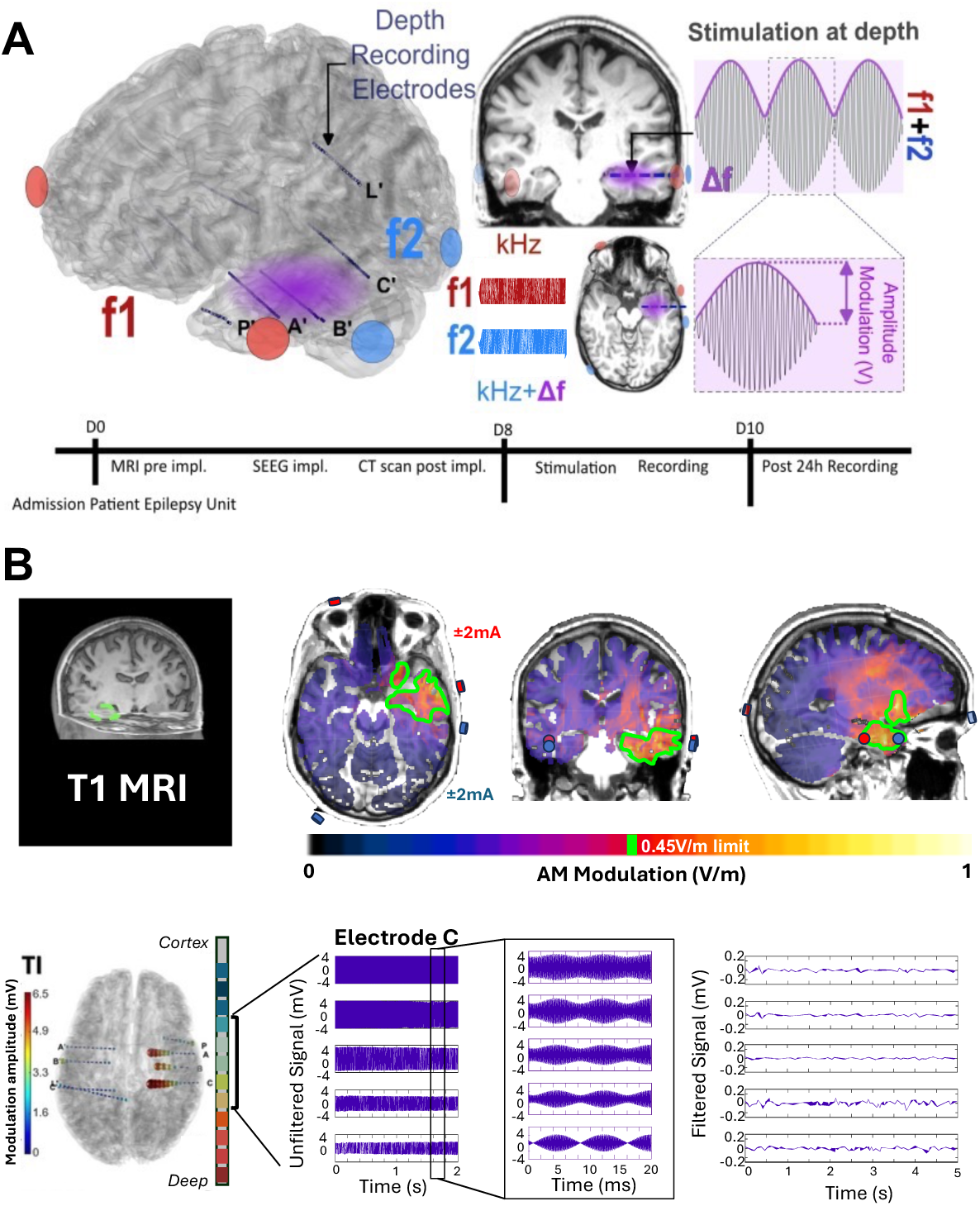
Temporal interference protocol in sEEG-implanted patients with epilepsy. (**A**) Scalp electrodes were positioned to deliver TI stimulation, consisting of two high-frequency currents which produce an AM field targeting the hippocampus in the temporal lobe. sEEG electrodes simultaneously record the electrophysiological signal and the modulated TI stimulation signal. Timeline of the experiments performed at the three centers: Admission Day 0, Stimulation Days 8-9. Each center applied TI with an AM frequency Δf = 130Hz, during a 20-minutes stimulation session and recorded the brain signal response after stimulation. (**B**) **Patient-specific head model and AM signal from TI stimulation.** MRIs of patients were used to create patient-specific head models using Sim4Life. (Top panel) the example simulation shows the placement of stimulation electrodes (red/blue) to target the ipsilateral hippocampus. The green border depicts an isocontour of the AM electric field, indicating the highest region of modulation centered on the hippocampus. (Bottom panel) simulations compare well to intracranial data, which similarly shows maximum AM in the temporal lobe (electrodes A, B, C), specifically in the hippocampus. Example recordings from the middle contacts of electrode C are shown. The raw signal (with using the average of all intracranial signals as reference) during TI stimulation allows visualization of the amplitude of the AM signal. When zooming in, the stimulation artifact revealed a well-defined AM in the deepest contacts, with diminishing magnitude approaching the cortex. The exact same recordings during TI stimulation are shown filtered with a bandpass filter ([1-1000] Hz) to extract electrophysiological signals. All oscillations below 500Hz were recovered, and reduced interictal spiking activity was observed for the TI condition.

### Biomarkers Detection

For the detection of biomarkers, electrophysiological signals were filtered (lowpass filtering [<1000Hz], performed using Matlab - MathWorks) to remove the stimulation artifacts. After down-sampling to 2500 Hz, semi-automatic detection of IEDs and HFOs (ripples and fast ripples) was performed using the AnyWave’s validated Delphos detector ^37^ – a well-established IED and HFO detector, regularly used in clinical epilepsy research^38^. Events markers (IEDs, HFOs) were extracted to determine their rates per minute. Subsequently, detected events were manually validated using AnyWave, a visualization software for electrophysiological data^37^.

### Statistics

The results of the detection process were imported into MATLAB (MathWorks), to facilitate comprehensive analysis and statistical assessment, and organized into distinct matrices based on patient identifiers, treatment centers (EMORY, INN-SU, or SAUH), and protocol conditions (baseline, stimulation, sham, post, and post 24H). From these matrices, initial visualizations were generated for each patient before aggregation for inter/intra-patient and inter/intra-center statistical assessments. IED, ripple ([80 250]Hz), and FRs ([250 500]Hz) rates were computed and juxtaposed across different stimulation protocols, as illustrated in Fig. 2, employing multivariate analysis of variance (ANOVA or Friedman test) and paired T or Mann-Whitney-Wilcoxon tests to discern significant variations. The aggregated dataset underwent further comparison via Friedman ANOVA and paired Wilcoxon tests to identify potential differences across all experimental conditions.

### Single neuron modeling

To study the interaction between a hippocampal CA1 pyramidal neuron and electrical fields we used a previously published model^39,40^, available online on the ModelDB database (a.n. 151731 and 190559, respectively). A detailed morphological and biophysical reconstruction of a CA1 pyramidal neuron^41^ (cellc62564 from Migliore *et al.* (2008), ModelDB a.n. 87535) was used for all simulations. A first set of simulations was performed in current clamp mode, injecting one or two currents for 1 sec and implemented as Equation 2 at different amplitudes *I*_0_.

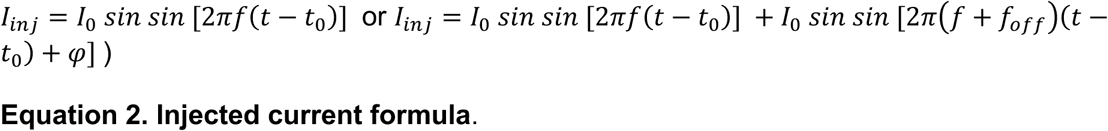

All simulations were performed using v.8.2.2 of the NEURON simulation environment (Hines and Carnevale, 1997)^42^.

## Results

### Temporal Interference Protocol in sEEG-implanted Epilepsy Patients

TI stimulation was performed in patients with drug-resistant mesial temporal lobe epilepsy implanted with sEEG electrodes (Figure 1A). Epilepsy patients with implanted sEEG electrodes offer a unique opportunity to precisely delineate the stimulated zone and assess the impact of TI exposure on brain activity. sEEG electrodes recorded the electrophysiological signal changes evoked by TI and mapped the AM exposure from the TI stimulation. The setup involved the placement of four scalp electrodes (2 pairs of 2 electrodes) to deliver TI stimulation, which consisted of two high-frequency signals unilaterally targeting the hippocampus (the side of the epileptogenic network) (Figure 1A and B).

### Simulation and Visualization of Stimulation Site

As seen in Figure 1B (top panel), an MRI image of a patient with stimulation electrodes specifically targeting the ipsilateral hippocampus is seen. The associated patient-specific simulation of the TI exposure targeting the hippocampus is visualized using Sim4life. The simulation estimates the electrode position for a correct stimulation of the hippocampus and delineates the region of strong AM modulation (green border) within the temporal region - with the maximum modulation located in the patient’s hippocampus.

### Recordings of AM Potential

An example sEEG recording of the AM potential during TI stimulation can be seen in Figure 1B (bottom panel). The amplitude of the AM was extracted and is depicted on the contacts of each sEEG electrode in the figure. The highest AM was observed on the electrodes within the temporal lobe, specifically in the hippocampus, and showed good agreement with the patient-specific Sim4Life simulations. The AM magnitude progressively diminishes towards more superficial cortical regions, while the magnitude of the carrier increases. An analysis of the filtered signal (1-1000 Hz) demonstrated the ability to extract electrophysiological signals (IEDs/HFOs), as TI stimulation artifacts are generally several thousand Hz higher than the electrophysiological signal of interest. No IEDs are visible in the example recording.

### Epileptic Biomarker Suppression with TI

All patients underwent baseline, stimulation, and post-stimulation recordings (see Figure 2A) consisting of 20 minute blocks: baseline recording, TI stimulation, post-stimulation recording (in the 20 minutes after the stimulation), and (at FNUSA only) an additional post-stimulation recording 24 hours after the stimulation session. During TI stimulation, patients did not report any symptoms, such as sensations associated with TI stimulation on the scalp, or other subjective symptoms with frequencies centered at 9kHz; 2 patients out of 3 receiving TI at 1/1.13kHz felt tingling on the scalp without any adverse events. There were no adverse events.

**Figure 2.**
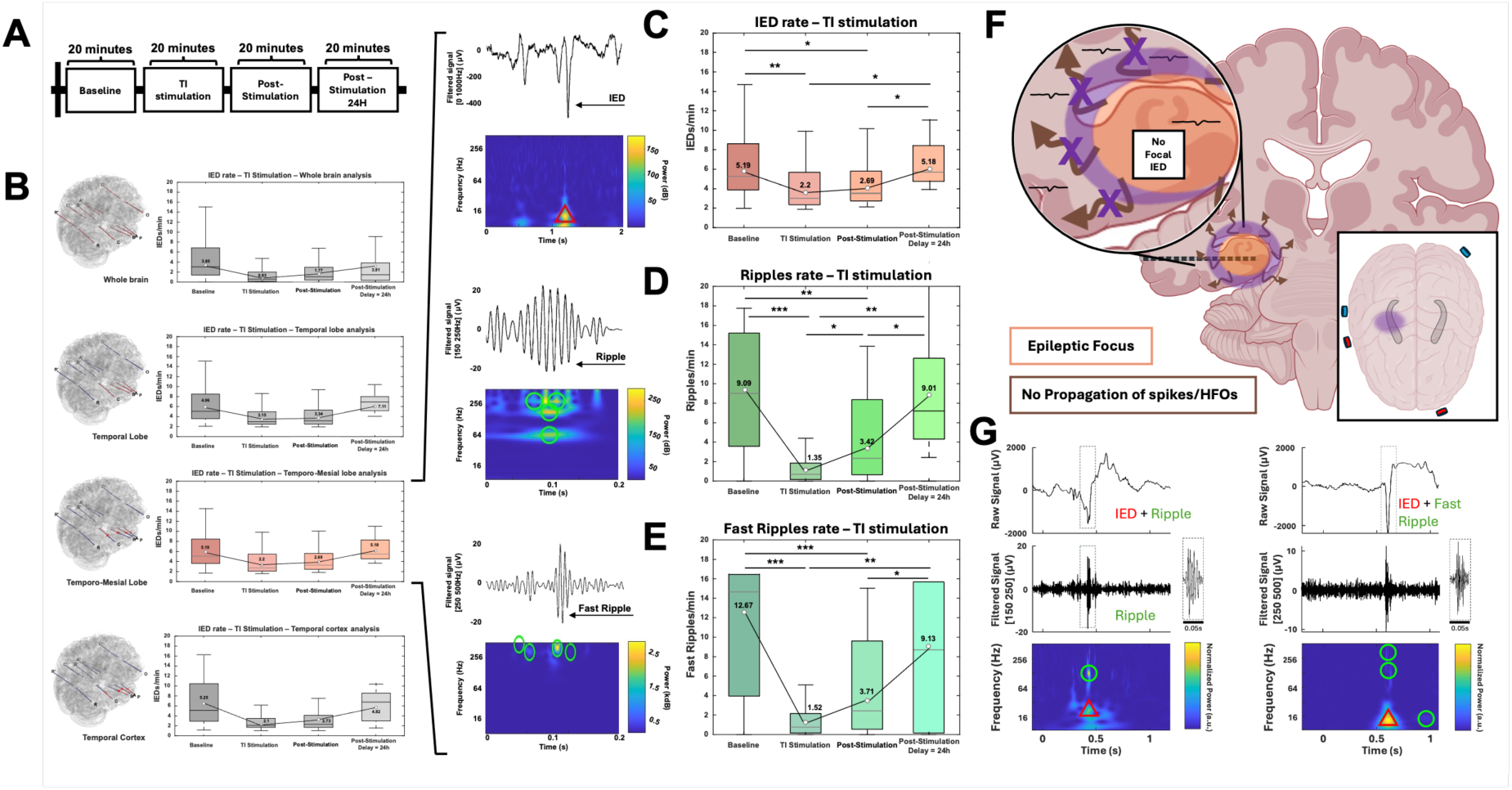
Epileptic biomarkers are suppressed during TI stimulation, and a post-stimulation carrier-over effect is observed. (**A**) The stimulation protocol was the same across the different centers (USA, Czech Republic, Hungary). Center-to-center differences are shown in Supplemental Figure S1. The protocol includes baseline recording (20 minutes), TI stimulation protocol (20 minutes; 30-second ramp-up and 30-second ramp-down included), post-stimulation recording (20 minutes), and – only in Czech Republic – post-24-hour recording (20 minutes). (**B**) Analysis of IEDs rates by brain region. All regions show a decrease in IED rate during TI stimulation. Analysis comparing all biomarker rates across all centers from the mesial temporal focus (n.s: p-value > 0.05; *: p-value ≤ 0.05; **: p-value ≤ 0.01; ***: p-value ≤ 0.001). (**C**, **D**, and **E**) Looking at the epileptic focus in detail: TI stimulation significantly decreases IEDs, ripples, and FRs, in a way similar to responses in DBS studies40,41. A feature of the TI biomarker suppression is that, in addition to the suppression during stimulation, the biomarkers do not return to their pre-stimulation values in the 20-minute period after stimulation – the suppression has a strong carrier-over effect. The 24-hour recordings indicate that the suppression is not permanent, seeing that biomarkers have returned to their pre-stimulation values. Brain-wide suppression of IEDs is a good indication that the focus of the epilepsy (the location of spike generation; in these patients the mesial temporal region, specifically the hippocampus) has been suppressed. (**F**) As the focus was suppressed, we expected and observe limited generation and therefore limited propagation of IEDs and HFOs. (**G**) The simultaneous suppression of HFOs and IEDs is understood via co-occurrence: it is well-known that HFOs strongly co-occur within IEDs^43,44^. This co-occurrence also appears in our data.

We compared the distribution of IEDs detected by sEEG in various brain regions (see Figure 2B): the whole brain (all SEEG contacts), the temporal lobe (electrodes A,B,C, and P from the most impaired side), the temporo-mesial area (deep contacts – 1 to 7 – from electrodes A,B,C, and P from the most impaired side)(the hippocampal focus), and the temporal cortex (shallow contacts – 8 to 15 – from electrodes A,B,C, and P from the most impaired side). All analyses were realized with the ‘global group’ encompassing all patients from the three centers. In baseline recordings, all patients showed signs of temporal lobe epilepsy, with a mesial focus. TI stimulation produced a statistically significant decrease in IEDs rate across all brain regions, which suggests that the focus was suppressed (IEDs originating from an unsuppressed additional focus would be visible). Post-stimulation data shows a strong carry-over effect with IEDs not returning immediately to baseline values. The post-stimulation recording 24 hours later shows that the IED rate returned to the baseline level, the carry-over effect is not permanent.

Specific analysis of TI’s impact on the epileptic biomarkers (IEDs, ripples, and FRs), specifically in the temporo-mesial region with the hippocampal focus, is presented in Figure 2C, D, and E. All epileptic biomarkers were statistically decreased during TI stimulation in the focus compared to the baseline recordings (Friedman ANOVA, p-value < 0.0001). In all three centers, TI stimulation of the mesial temporal lobe was correlated with a decrease in IEDs (data for each participating center in supplemental figures). The relative decrease ranged up to 86.3% (58.5 +/− 27.8%, IC95%) depending on the patient (Friedman ANOVA, p-value_TI stimulation vs. baseline_ < 0.0001).

For HFOs, it is well established that excessive pathological ripples^43^ and pathological fast ripples^44^ co-occur within IEDs (see Figure 2G). Our data shows a reduction of IEDs with a corresponding reduction of co-occurring HFOs^45,46^.

For all three centers, the stimulation effect was sustained, as evidenced by a significant decrease in IEDs and HFOs 20 minutes after stimulation (Wilcoxon tests, p-value_Post-stimulation 20 minutes vs. baseline_ < 0.0001). TI stimulation (Figure 2F; purple) targeting the pathological mesial focus (orange) shows a strong suppression of epileptic biomarkers, and a suppression of the propagation of biomarkers, with a sustained post-stimulation carry-over effect.

### Epileptic Biomarkers Suppression with kHz (Sham) Stimulation

As TI is delivered via a combination of kHz frequencies, we investigated the effect of sham stimulation, where both pairs of electrodes provide the same frequency, and no offset is present between f1 and f2 (Figure 3A).

**Figure 3.**
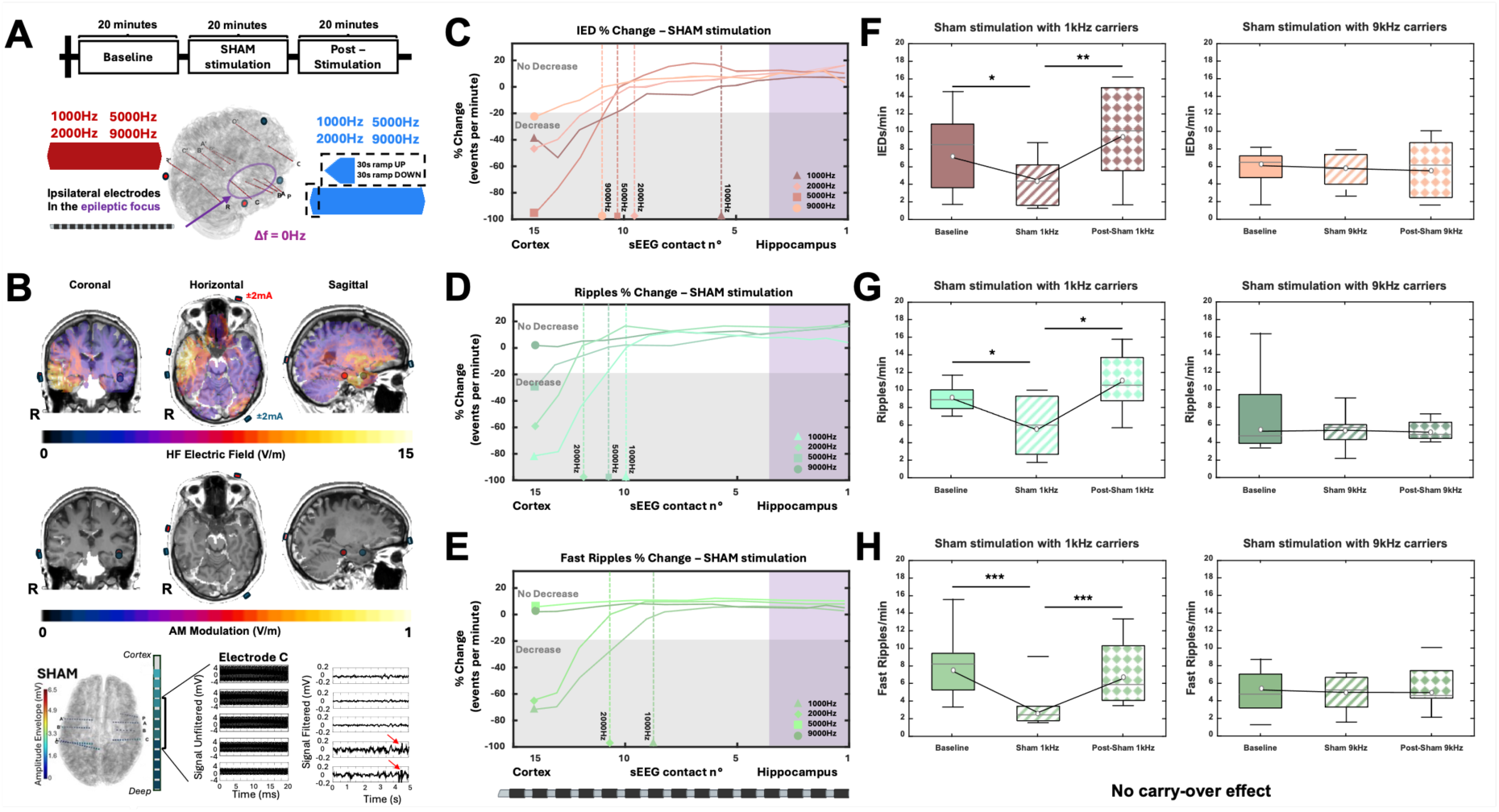
Sham stimulation and HFO suppression. Sham stimulation consisted of applying the same frequency (f_1_ = f_2_) to both pairs of electrodes. The protocol is 20 minutes baseline recording, 20 minutes sham stimulation and recording, and 20 minutes post-stimulation recording. A 30-second ramp-up and a 30-second ramp-down is included to avoid unwanted transients. The stimulation amplitude is ±2mA (4mA peak-to-peak) (**A**). As for the TI stimulation condition, a patient-specific simulation is performed to estimate the kHz field strength and illustrate the absence of AM. The sham voltage recorded by sEEG in a patient is visualized. After filtering, example IEDs (red arrows) are apparent in the hippocampus recordings, but not in the cortical ones (**B**). The changes in biomarker rate between baseline and sham stimulation are plotted as a function of location along the ipsilateral sEEG electrode entering the pathological hippocampus. The zero line (dashed line) and grey region denote the absence of a significant rate difference between baseline and sham. sEEG contact locations in the epileptic focus are highlighted in purple. A biomarker rate reduction of over 20% is considered a clear indication of sham suppression. As can be seen, suppression penetrates the deepest at 1kHz. At 9kHz change remains below ±20% throughout for all biomarkers (**C**, **D**, **E**). Biomarkers are plotted for the three parts of the sham protocol using the lateral cortical contacts – sEEG contacts 15 to 8 (**F**, **G**, **H**). 1 kHz caused a significant suppression during stimulation, potentially due to conduction block. 9 kHz did not cause significant suppression. In each case, there was no visible carrier-over effect, unlike in the TI conditions.

As seen in Figure 3A, all sham patients underwent the same protocol in blocks of 20 minutes: a baseline recording, kHz sham stimulation (two pairs of electrodes with no offset frequency – applying the same currents as for TI stimulation), and a post-stimulation recording (for 20 minutes after stimulation). The electrode locations were also identical to the TI stimulation electrode locations. Patient-specific Sim4Life simulations were performed to determine the carrier field magnitude distribution. The illustrative visualization in Figure 3B delineates the region of maximum kHz exposure (top panel), situated in the temporal region, and the expected absence of AM (middle panel). sEEG electrodes were used to record the electrophysiological signal changes evoked by sham and to map the sham exposure potential (bottom panel). The example recording features IEDs in the mesial focus (highlighted by red arrow), but not in the cortex where there is a higher kHz field magnitude.

The effects of kHz sham on biomarker rates are shown in Figure 3C, D, and E, as a function of depth along the sEEG electrode into the ipsilateral hippocampal focus. IEDs, ripples, and FRs rate changes are pictured in C, D, and E respectively. Fluctuations outside of the ±20% range are considered a clear indication of kHz suppression of a biomarker, rather than physiological variations between the two 20-minute sessions of baseline and sham^47^. Vertical dotted lines represent the depth at which the sham stimulation starts to decrease the biomarkers. For the considered electrodes, contacts #15 – #10 are located in lateral cortices and contacts #9 – #1 are in the medial cortices (including hippocampus).

Higher frequencies (i.e. 7 and 9 kHz) show less biomarker reduction compared to lower carrier frequencies (i.e. 1 and 2 kHz) in the superficial cortices. Lower frequencies are, furthermore, associated with deeper penetration of rate changes. 9kHz sham minimally impacts these biomarkers.

As shown in detail for 1kHz and 9kHz in Figure 3 F, G, and H, a common trend is visible across all biomarkers: sham-stimulation induced decreases in IEDs and HFOs (ripples and fast ripples) at lower kHz frequencies in the lateral, but not deeper medial cortices/hippocampus, and had no significant decrease at 9kHz. Additionally, for low-frequency kHz biomarker suppression, there is no measurable carry-over effect.

Previous studies using simulations of injected charge with a Hodgkin-Huxley (HH) axon model have suggested kHz conduction block as a possible side-effect of TI deep brain modulation^48^. Specifically, models predict that a deep brain region may respond to the AM signal, but shallower brain regions will be subjected to stronger kHz fields which may lead to conduction block. Although perhaps not “conduction block” in the traditional sense of suprathreshold stimulation of peripheral nerves, we believe that we have observed a similar phenomenon when using lower frequency kHz carriers for TI, and as predicted by the models, the phenomenon diminishes as the kHz frequencies increase further - as neural activation thresholds increase with kHz frequency (Figure 4B). To avoid the phenomenon, one can use a higher carrier frequency. However, some models predict that shifting the carrier frequency up (while maintaining Δf constant) to reduce conduction block, will simultaneously reduce the modulatory effectiveness of AM. We do not observe this experimentally. More precisely (deep contacts – 1 to 7 – from electrodes A, B, C, and P from the most impaired side), we did not see any reduction in the effectiveness of epileptic biomarker suppression by using higher carrier frequencies when comparing the biomarker rates during TI stimulation (Wilcoxon tests, p-value_TI1000Hz vs TI9000Hz_ > 0.05).

**Figure 4.**
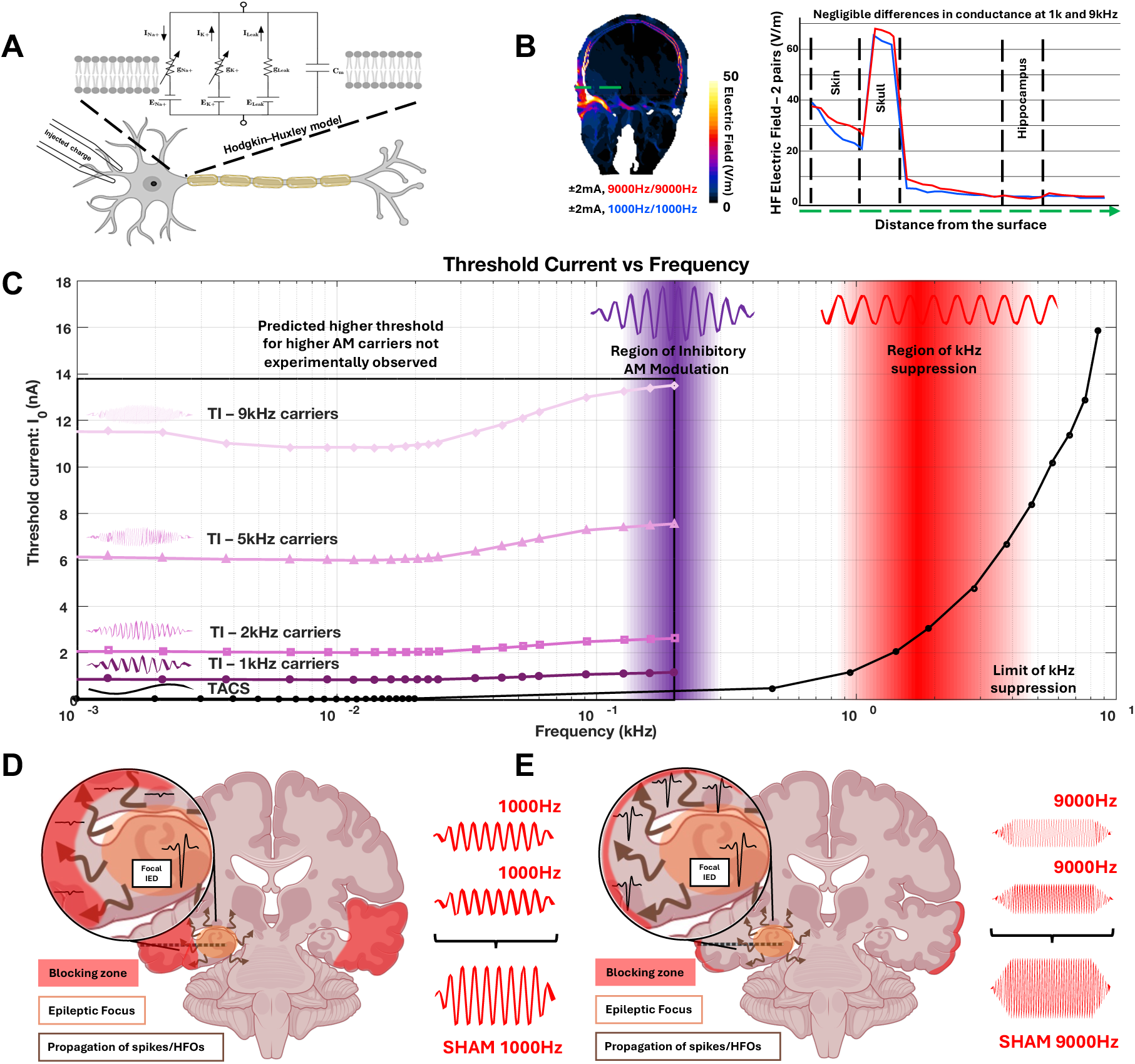
Biophysics Insights. **A)** Previous work simulating exposure-induced suprathreshold neuromodulation of a Hodgkin–Huxley-like (HH) axon model has highlighted the potential for conduction block in TI stimulation^48^. Our sham stimulation results are consistent with some of the concerns raised by those authors. **B)** Conductivities and permittivities implemented in the simulations are based on Gabriel et al., 2009^49^; values can be revised if new conductance measures become available. The carrier frequency dependence of sham effects cannot be explained by field magnitude differences, as electrical conductivity hardly depends on frequency below 10 kHz, such that for identical channel current magnitudes the quasistatic solutions to Maxwell equations are very similar – only at much higher frequencies does the frequency dependence of conductivity and its impact on displacement currents become relevant. Also head and contact impedance changes as a function of frequency are fully compensated for by the current control of the sources (see Supplemental Figure S5). **C)** Experimental work by Bernard Katz^50^ and simulations with an HH-like neuron model, showed that the threshold for propagation blocking by unmodulated exposure (red shading) increases with frequency (the threshold obviously depends on diameter, fiber type, etc). This is in accordance with the kHz suppression (where we use the word suppression and not block, as we apply subthreshold electric fields) of IED propagation from the mesial focus observed in this study for low kHz sham frequencies and the experimentally demonstrated decrease in the suppression depth with increasing carrier frequency (fading red). However, the HH model fails to replicate the experimentally observed absence of a strong carrier frequency impact on AM effectivity. Moving to a higher carrier frequency eliminated sham effects - however, the AM signal created with the same higher carrier frequency suppressed the epileptic focus without apparent reduction in efficacy (purple region). **(D – E).** Support for assuming that two different mechanisms are at work could be derived from the lack of a carry-over effect from sham stimulation (turning off the high frequency allowed activity to propagate normally again), while AM exposure produced a strong carry-over effect, where suppression of the hippocampal focus resulted in a continued reduction of epileptic biomarkers post-stimulation.

Overall, these results suggest that TI stimulation can reduce HFOs and IEDs, when targeting the hippocampus with a modulation at a traditionally inhibitory frequency (130 Hz). For sham, mid/low-carrier frequencies (≤ 5kHz) also decrease the number of epileptic biomarkers, but only in superficial cortices. With a 130 Hz AM, the highest carrier frequency we tested (9 kHz) caused a significant decrease in all epileptogenic biomarkers, while the corresponding sham condition, did not change ripple and fast ripple rates, and had the least effect on IEDs. The carrier frequency selection seems to play an important role for the use of TI as a focal, non-invasive form of deep brain modulation, i.e. 9kHz induces less off-target neuromodulation than 1kHz, due to the absence of kHz carrier field effects.

## Discussion

Here we show for the first time that a non-invasive deep brain modulation method, Temporal Interference, can selectively target the epileptic focus and reduce epileptiform activity in epilepsy patients, as verified by intracerebral EEG recordings. We show that TI can create an AM signal at depth in the human mesial lobe and that TI stimulation at Δf = 130Hz can significantly reduce IEDs, ripples, and FRs during stimulation. Furthermore, we show that the choice of carrier frequencies matters for TI stimulation: As illustrated in Figure 4A and C, our results are consistent with the presence of biomarker suppression, primarily in superficial structures, at low kHz frequencies. However, suppression mechanisms explained by high-frequency conduction block, as well as simple HH models of neural dynamics, fail to explain how TI with higher kHz carriers produces targeted, non-invasive, deep brain modulation, with no penalty in terms of effectiveness compared to TI with lower kHz carriers – something that previous studies had ruled out based on theoretical and computational considerations^51^. This highlights the importance of clarifying the mechanisms underlying TI neuromodulation.

In terms of future utilization in epilepsy, the application of TI stimulation represents a significant advancement in the pursuit of non-invasive diagnosis and therapy for epilepsy, particularly for patients who are not suitable candidates for resective surgery. Unlike conventional transcranial electrical stimulation techniques, such as tDCS and tACS, which are limited by the dominance of superficial cortical effects, TI stimulation allows for the modulation of neural activity in deeper brain structures without invasive procedures. This is particularly relevant for targeting epileptogenic zones located in areas of the brain which are not easily accessible by traditional stimulation methods or only by electrical stimulation via depth probes^52,53^.

In previous work, TI stimulation in the peripheral^29^ and central nervous systems modulates population-wide neural activity^54^ as well as individual neuronal activity^55^, when employing Δf frequencies similar to those found to be effective when applied directly (i.e. Δf = 1Hz modulating activity at a rate of 1 Hz – as is expected if 1Hz is directly applied). Our findings here are similarly in line with previous research, namely studies conducted in the hippocampus of mice in an epilepsy model^27^ as well as with various clinical studies which have used DBS stimulation (130-145Hz) in the hippocampus of patients to manage their seizures and decrease epileptogenic biomarkers^3,10,13,14,56–58^.

Future work could leverage the ability of TI to reduce IEDs non-invasively and in a targeted manner, as there is growing awareness that IEDs can transiently disrupt focal and global cognitive processes, and anti-seizure medications are currently not deliverable in a focal manner^59–62^. In that sense, TI could be a valuable tool in neurological investigation of cognitive task performance with epilepsy patients. Indeed, even for conduction block, as we observed for lower kHz frequencies, it is possible that this mechanism could be used beneficially in the context of epilepsy in future work.

While the presented results are promising, several limitations must be acknowledged. First, the study’s reliance on patients already implanted with sEEG electrodes for epilepsy monitoring and who feature identifiable seizure foci means that the findings may not be directly generalizable to the broader population with refractory epilepsy – for example in generalized seizures, where identification of specific EZs is not possible. Furthermore, we only investigated medial temporal/ hippocampal and not neocortical EZs, because we aimed at demonstrating the feasibility of stimulating deep structures, and for the purpose of acquiring sufficient patients with a similar target given that this is the most common type of epilepsy in adults. Additionally, the specificity and efficacy of TI stimulation in reducing seizures and seizure frequency remains to be fully established through longitudinal studies with larger sample populations, but the benefit of other neuromodulatory therapies correlates with IED suppression.

## Conclusion

In conclusion, the application of TI stimulation in patients with epilepsy represents a novel and promising approach to the non-invasive treatment of epilepsy. While preliminary findings are encouraging, further research is essential to elucidate the potential of TI stimulation to help manage focal epilepsy, in particular as a tool to assess neurostimulation responses prior to DBS or RNS implantation.

Future research should expand the number and diversity of studied patients undergoing TI stimulation, to better understand its efficacy and safety profile across different types of epilepsy and to assess the long-term impact of stimulation. Additionally, further investigation into the mechanisms underlying the effects of TI stimulation are required to enhance our understanding of TI and epilepsy pathophysiology and to be able to systematically optimize and personalize stimulation parameters in the spirit of precision medicine.

## Acknowledgments

A.W. received funding from the European Union’s Horizon Europe research and innovation programme under grant agreement No. 101101040 (TREATMENT) and No. 101088623 (EMUNITI). Efforts on this project for both D.L.D. and E.A. were in part supported by grants received from the National institute of Neurological Disorders and Stroke (NINDS) of the National Institutes of Health (NIH) [R01NS088748]. E.A. was also supported through the Emory Neuromodulation Technology Innovation Center (ENTICe) and a Catalyst Award from the Emory Neurosurgery Department to C.A.G. and D.L.D. E.A. was also supported by the American Epilepsy Society (AES) post-doctoral fellowship. We thank S. Dabiri for his assistance in downloading MRI and electrophysiological recordings. We extend our thanks to the dedicated nurses at the epilepsy units of Emory University, St. Anne’s University Hospital, and Semmelweis University for their invaluable support. This work received financial support from Agence Nationale de la Recherche under France 2030 bearing the reference “ANR-24-RRII-0005 », on funds administered by Inserm.

## Authors Contribution

A.W. conceived and designed the project. F.M., E.A., A.D., J.T., O.S., C.L., M.A.S., E.B., V.V., J.S., D.F., conducted human experiments and acquired neural data. F.M. and E.A. analyzed neural data. F.M. created the patient-specific head models and performed the associated finite-element models. R.M. and M.M. designed and analyzed single-neuron models. F.K. and E.N. investigated the implant related field enhancement. F.M., E.A., D.L.D., and A.W. wrote the first draft of the manuscript and refined it after inputs from I.D., M.P., C.A.G., S.L., R.C., F.K., R.C.A., A.M.C., N.K., E.N., F.B., N.P.P, R.E.G, V.J., and M.B.

## Data Availability Statement

All data produced in the present study are available upon request to the authors.

## Supplementary Figures

**Figure S1.**
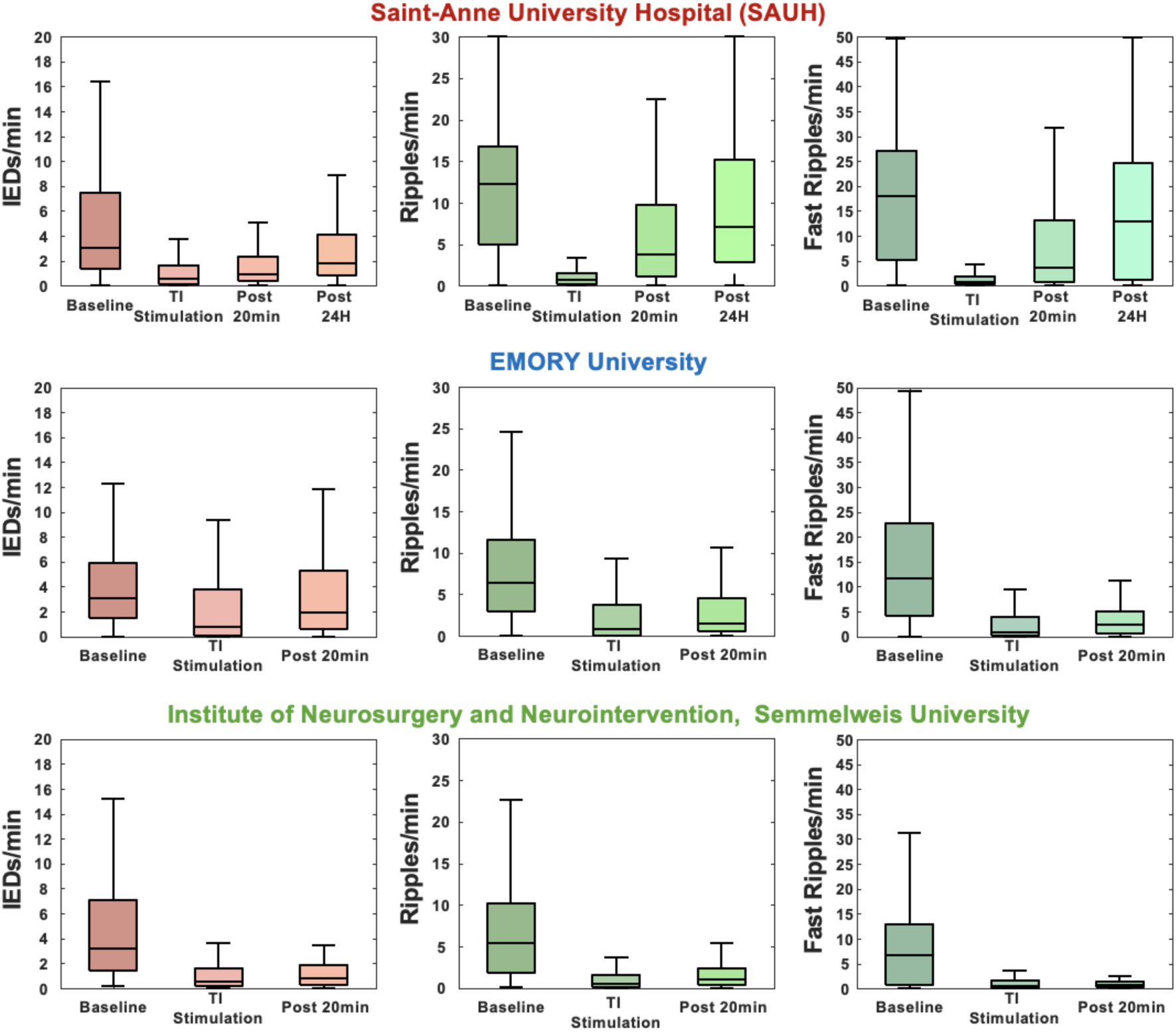
Center-by-center analysis.

**Figure S2.**
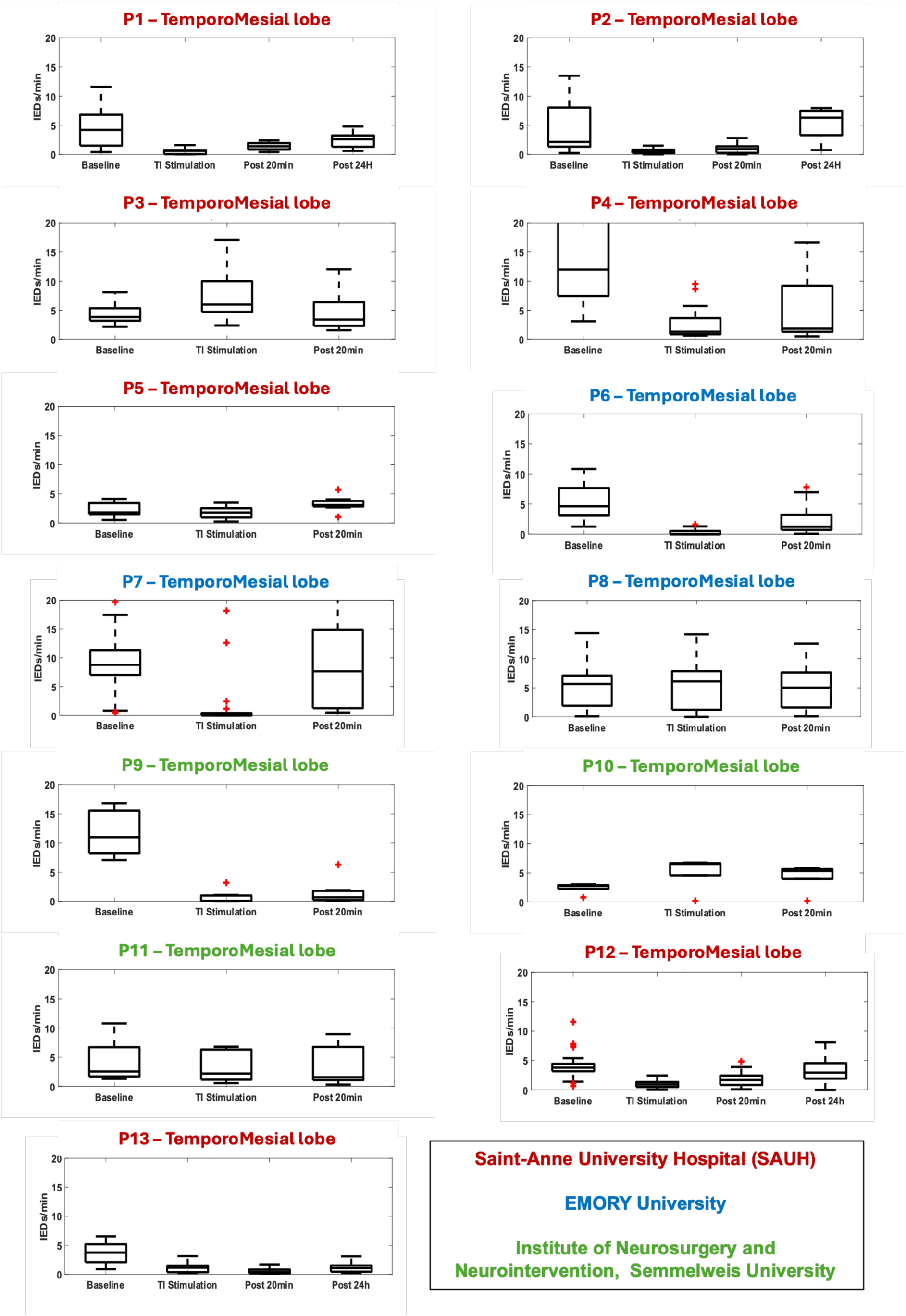
Single patient interictal epileptiform discharge analysis.

**Figure S3.**
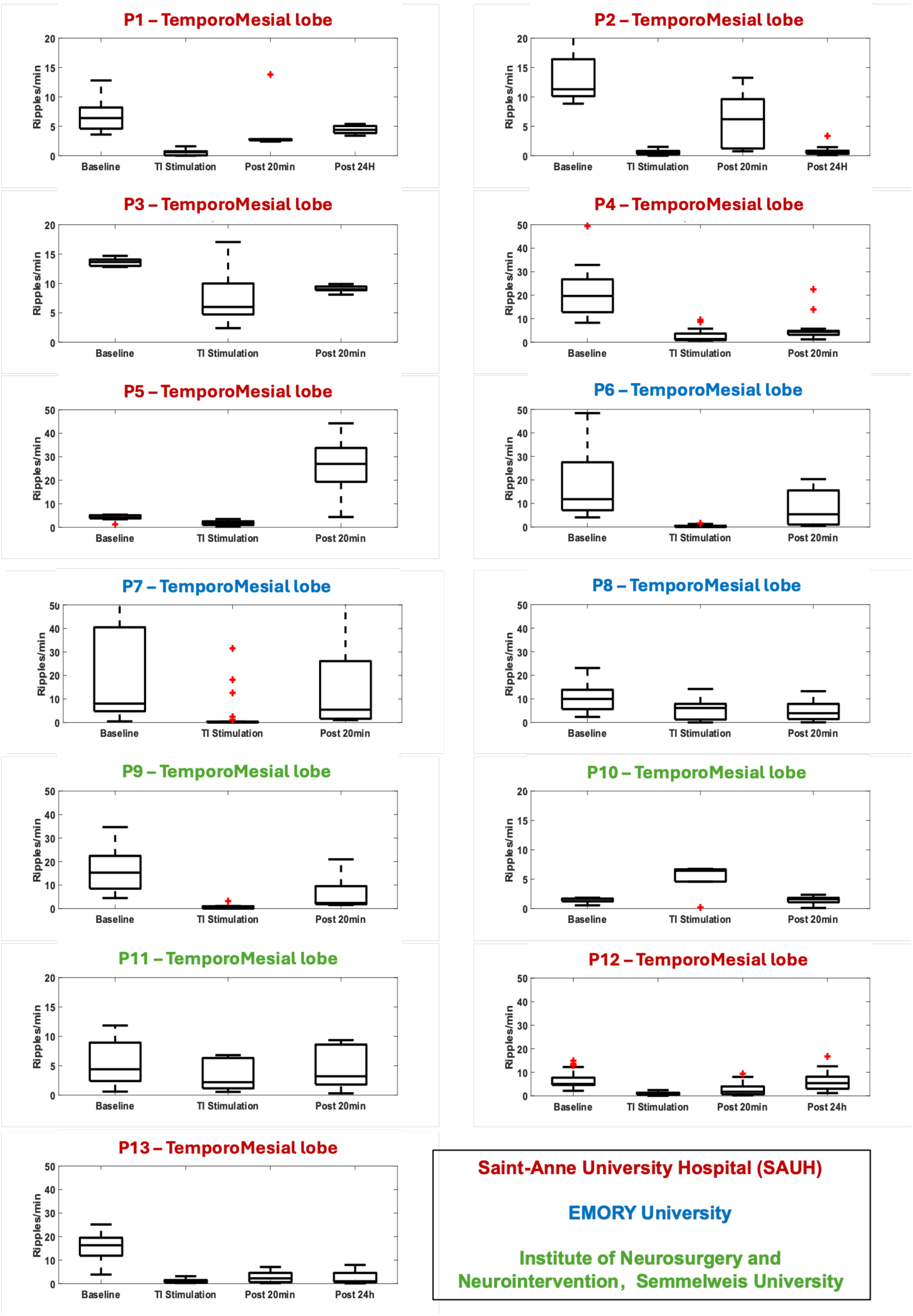
Single patient ripples analysis.

**Figure S4.**
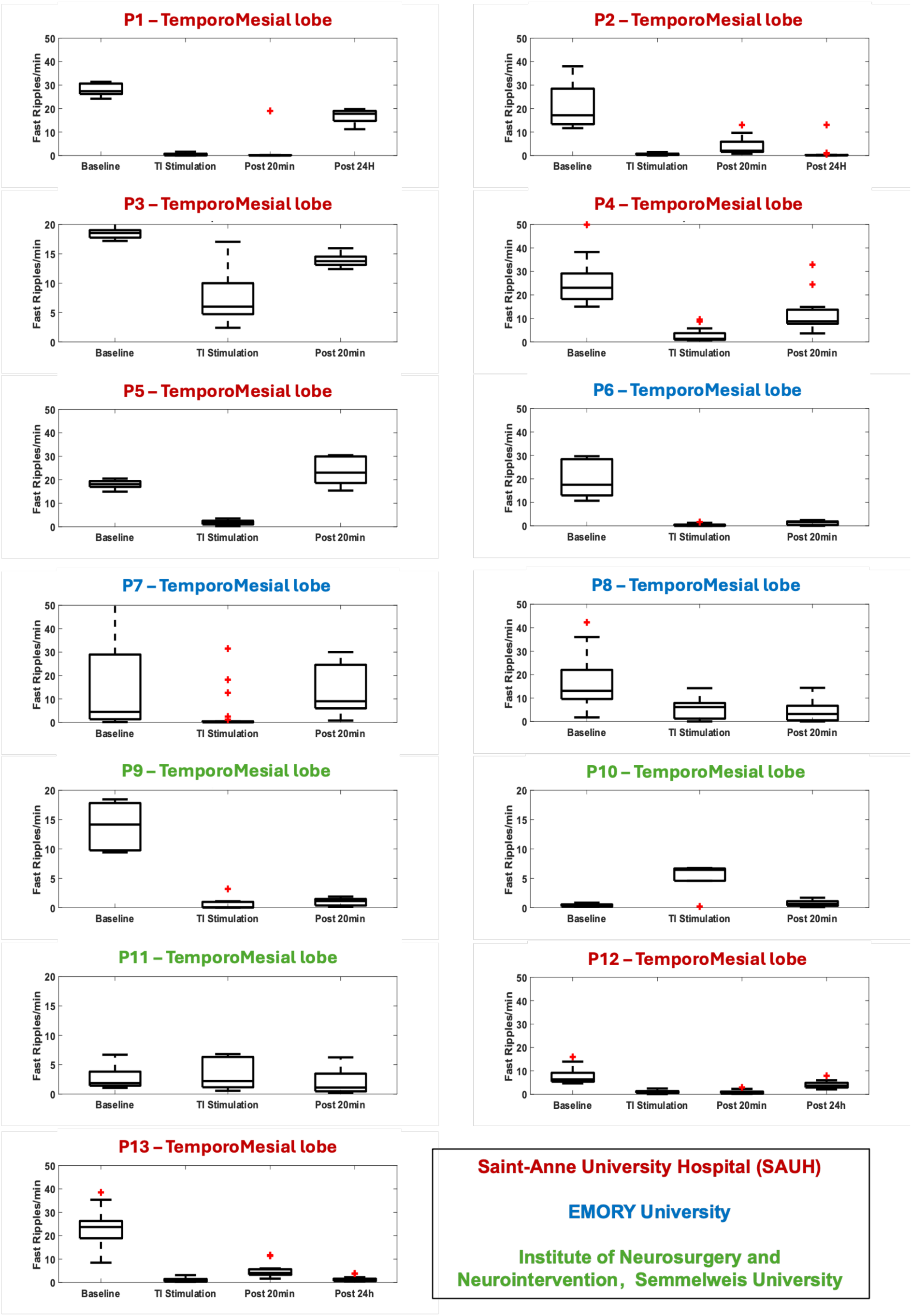
Single patient fast ripples analysis.

**Figure S5.**
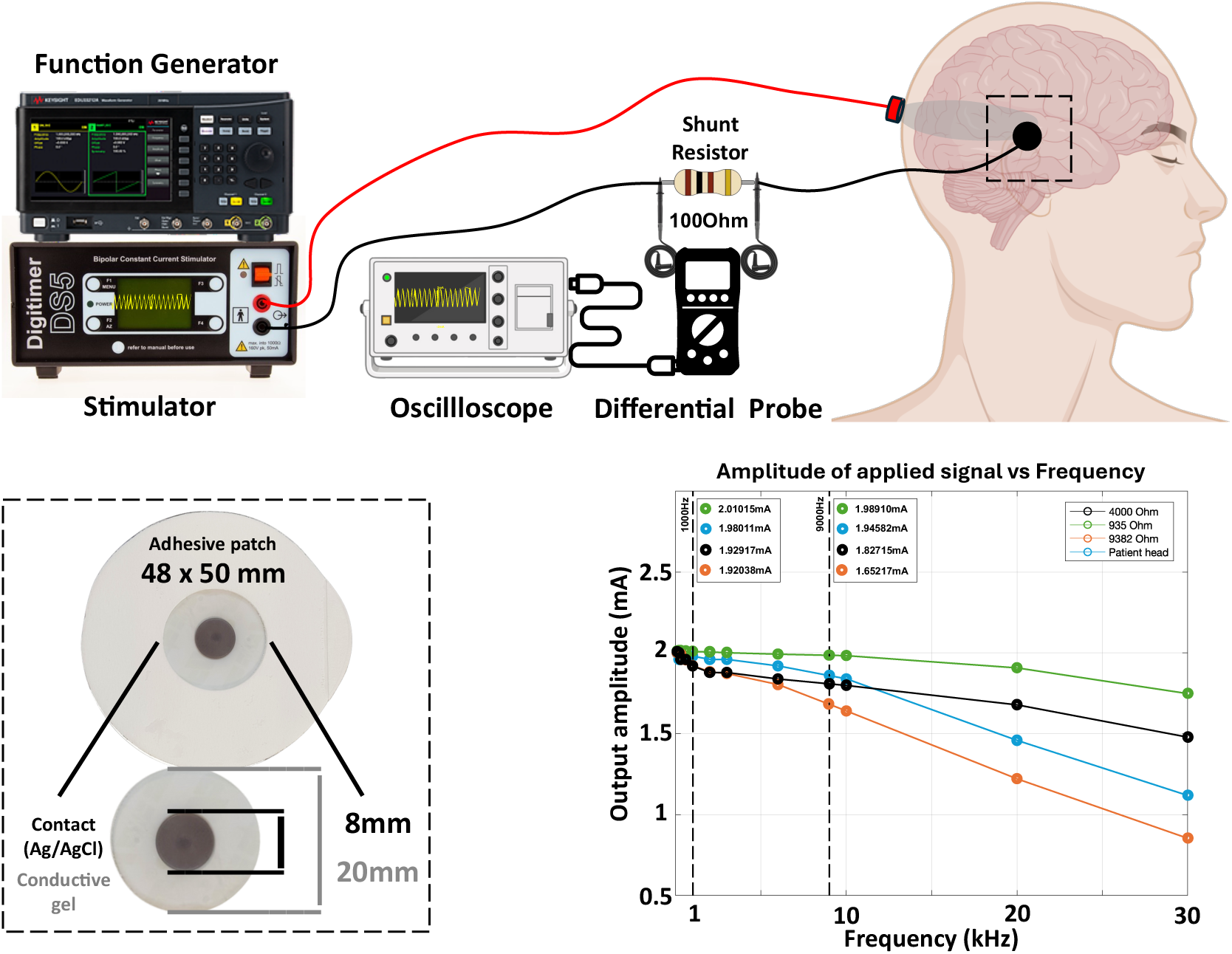
Simulations of the TI stimulation for different carrier frequencies. Measurement across a 100Ohm resistor of the output current as a function of the frequency applied via the Keysight and DS5. The relationship shows no significant applied current difference between lower carrier values (1000Hz) and higher carrier values (9000Hz) for a biological impedance (patient head – blue trace), effectively demonstrating the devices show no frequency roll-off effects from the spectrum of carrier values and impedances that were used in the study.

**Figure S6.**
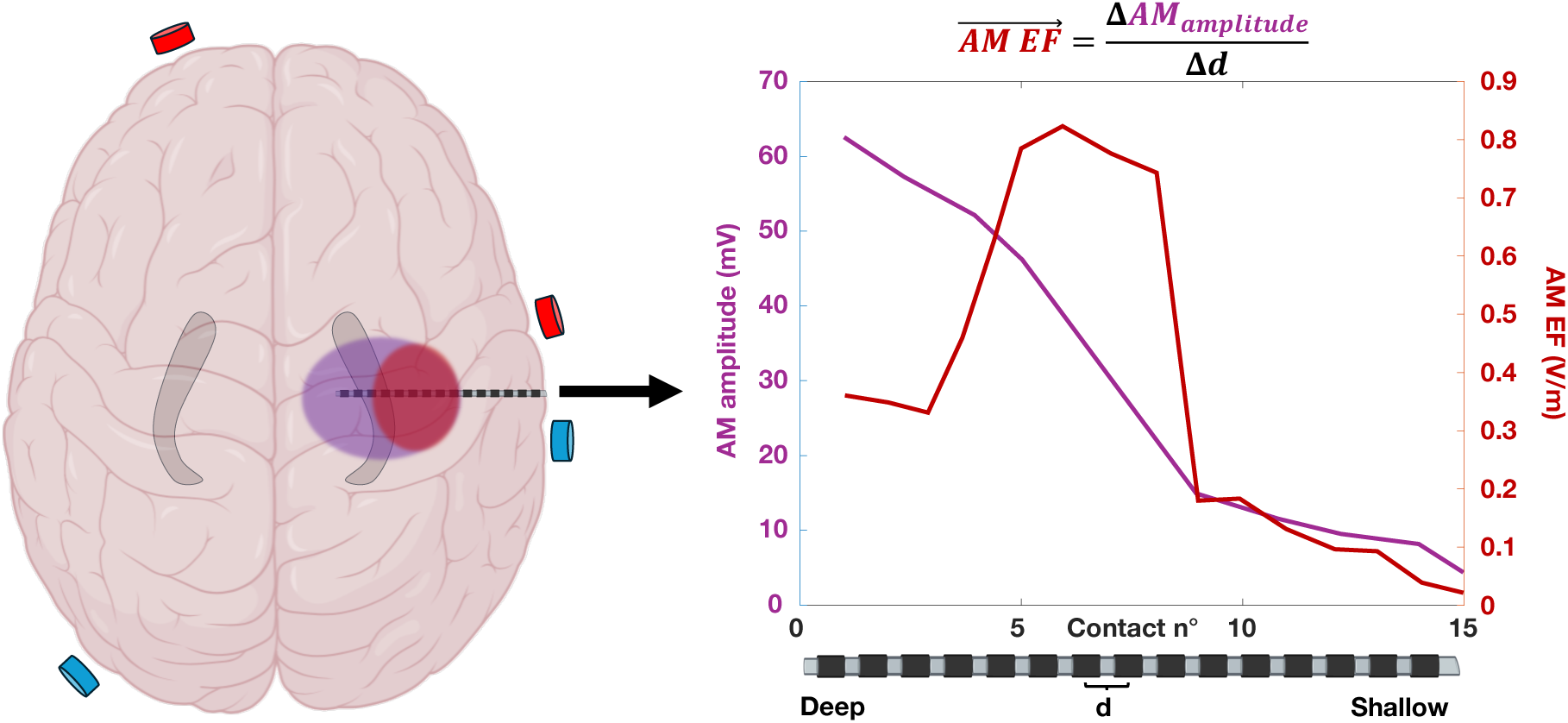
Recorded AM amplitude and calculated AM EF along an sEEG electrode targeting the hippocampus. The distribution of the maximum modulation amplitude and the maximum modulation electric field will differ. (**right panel**) The experimentally measured maximum of the AM corresponds well to the maximum of the AM electric field (computed as the modulation magnitude of the difference of potential between two contacts) at the focus of the epilepsy in the hippocampus. (**right panel**) The source of the neuromodulation effect is assumed to be the maximum AM field exposure.

**Figure S7.**
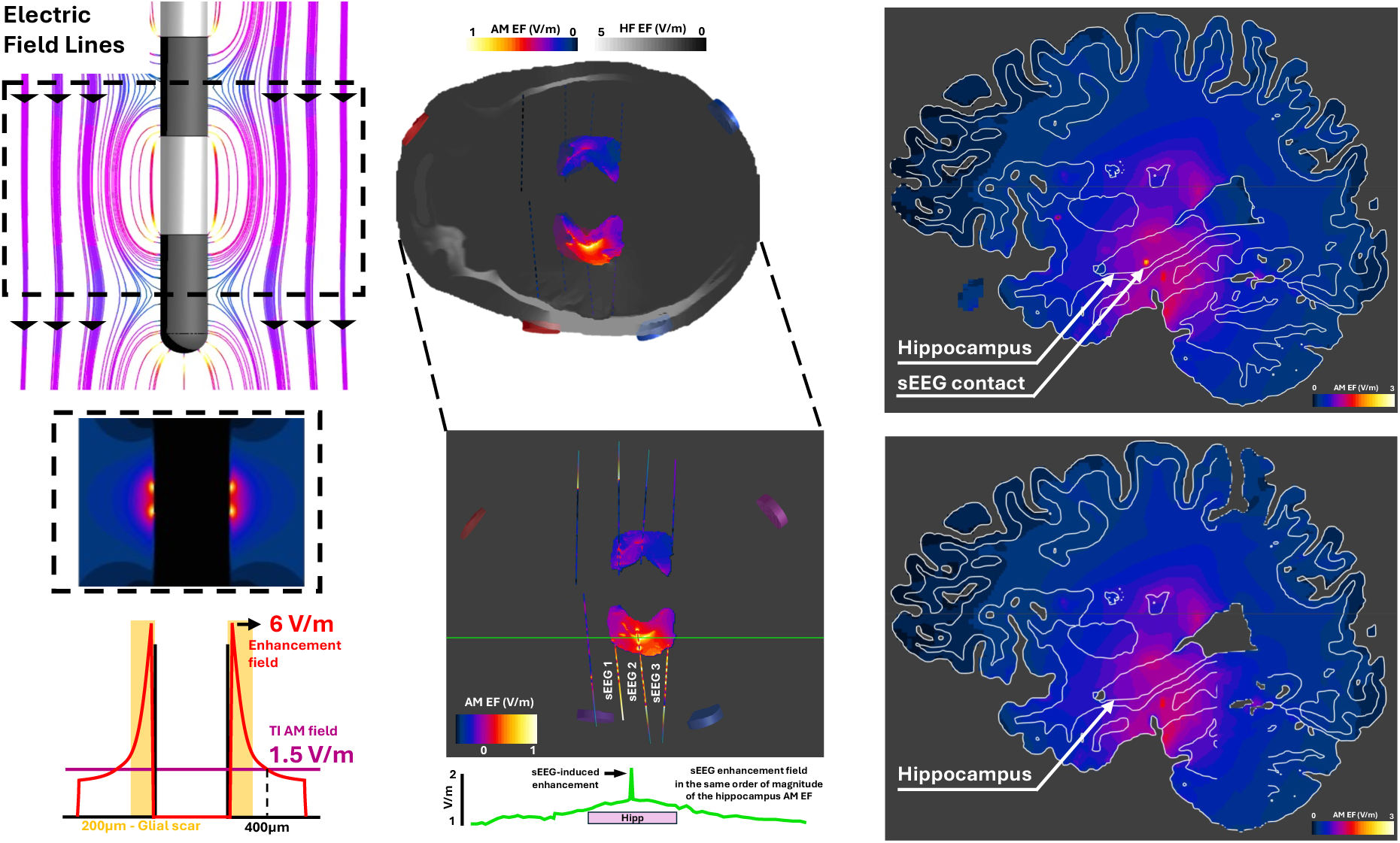
Theoretical Field Enhancement around sEEG electrodes. We characterize the electric field enhancement around the implanted SEEG electrodes during transcranial stimulation and determine the expected impact to be negligible. **(left panel - top)** Currents injected by transcranial stimulation can leverage low-impedance pathways through metallic SEEG contacts, and field lines bend near metallic surfaces such that tangential field components vanish. This not only results in local field enhancement^51^ near contact edges and bridging currents between neighboring contacts, but also in alignment of the two TI fields, resulting in an associated modulation magnitude increase. **(left panel – bottom)** The enhancement of the field was computed and found to be 2 to 7.2x the size of the incident TI modulation magnitude but decayed to the level of the surrounding AM field within 400 µm. Glial sheath formation around implanted intracranial electrodes is estimated to range from 150 to 300 µm. **(middle panels)** Visualization of the enhancement in a patient model – horizonal slice: the TI field near SEEG contacts in the hippocampus is depicted and plotted along the green trajectory. Several highly localized spots of AM enhancement are apparent. **(right panels)** Similar enhancement is evident in a sagittal cross-section. As can be seen in the middle and right panels, the entire hippocampus (left side) is covered by high AM exposure. The suppression of epileptic biomarkers at the brain network level, as observed in the current study, is unlikely to be caused by the highly localized subthreshold modulation near electrode contacts, considering that no significant enhancement is present outside the typical glial sheath thickness.

**Table 1.** Participant characteristics. Ages are displayed in ranges of 5 years to ensure anonymization of the patients.

| Patient | Age | Sex | Center | Head Ø | N° of electrodes | SOZ | Day PI |
| --- | --- | --- | --- | --- | --- | --- | --- |
| P1 | 35-39 | F | SAUH | 59 | 8 | Right temporal lobe | 6 |
| P2 | 40-44 | M | SAUH | 61 | 13 | Left Insular lobe | 6 |
| P3 | 35-39 | M | SAUH | 60 | 11 | Left temporal lobe | 6 |
| P4 | 45-49 | F | SAUH | 60 | 8 | Bi-temporal lobes | 6 |
| P5 | 40-44 | M | SAUH | 61 | 8 | Left temporal lobe | 6 |
| P6 | 35-39 | M | EMORY | 54 | 10 | Bi temporal lobe | 7 |
| P7 | 25-29 | M | EMORY | 60 | 13 | Left parietal lobe | 7 |
| P8 | 35-39 | M | EMORY | 58 | 18 | Left temporal lobe | 7 |
| P9 | 30-34 | F | INN-SU | 58 | 8 | Left temporal lobe | 8 |
| P10 | 40-44 | M | INN-SU | 61 | 6 | Left temporal lobe | 7 |
| P11 | 45-49 | F | INN-SU | 60 | 7 | Right temporal lobe | 6 |
| P12 | 25-29 | M | SAUH | 61 | 12 | Right temporal lobe | 6 |
| P13 | 30-34 | F | SAUH | 60 | 14 | Left temporal lobe | 6 |

